# Safety and Immunogenicity of 20-Valent PCV According to Number and Timing of Primary Series Doses

**DOI:** 10.64898/2025.12.23.25342907

**Authors:** Mary Kline, Anne L. Wyllie, Kathleen McElwee, James Trammel, Ayman Sabra, Jennifer C Moïsi, Christian Theilacker, Calman A. MacLennan

**Affiliations:** Vaccine Research and Development, Pfizer Vaccines, Pfizer Inc, Collegeville, PA 19426, USA; Pfizer Vaccines, Pfizer Inc, Cambridge, MA 02139, USA; Pfizer Vaccines, Pfizer, Paris, France; Pfizer Vaccines, Pfizer Pharma GmbH, Berlin, Germany; Vaccine Research and Development, Pfizer Ltd, Marlow, UK

**Keywords:** infants, immunogenicity and safety, 20-valent pneumococcal conjugate vaccine, *Streptococcus pneumoniae*, clinical trial, dosing interval, timing, 2+1, 3+1

## Abstract

**Background:** The number and timing of pneumococcal conjugate vaccine (PCV) primary doses can impact infant immune responses. This descriptive post hoc analysis evaluates the immunogenicity of the 20-valent PCV (PCV20) by vaccination timing in healthy infants in 2 key phase 3 trials.

**Methods:** Immunogenicity endpoints from study B7471012 comparing PCV20 to 13-valent PCV (PCV13) in a 2+1 schedule were examined by timing of vaccination subgroup; the 2,4m subgroup (participants vaccinated at 2, 4, and 11–12 months of age), and the 3,5m subgroup (participants vaccinated at 3, 5, and 11–12 months). Additionally, immune responses from the 2+1 schedule were compared descriptively with those in study B7471011, in which participants received PCV20 or PCV13 in a 3+1 schedule at ∼2, 4, 6, and 12−15 months.

**Results:** Serotype-specific serum immunoglobulin G (IgG) responses in the PCV20 3,5m subgroup were higher than those in the 2,4m subgroup across all 20 vaccine serotypes and comparable to PCV13 immune responses in the 2,4m subgroup. Regardless of primary doses timing, PCV20 2+1 elicited strong immune responses after the toddler dose leading to post-toddler dose IgG levels similar to a 3+1 schedule. PCV20 was safe and well tolerated in both the 2,4m and 3,5 m subgroups.

**Conclusions:** In this descriptive analysis of the 2+1 schedule, a 1-month delay in PCV20 priming to 3 and 5 months was associated with improved immune responses comparable with those of PCV13 at 2 and 4 months. NCT04546425; NCT04382326.

## INTRODUCTION

*Streptococcus pneumoniae* is an important cause of morbidity and mortality globally in infants and children <5 years of age.^1^ Widespread pneumococcal conjugate vaccine (PCV) use has led to substantial decreases in pediatric pneumococcal disease.^2^ It is estimated that from 2010‒2019, the 13-valent PCV (PCV13) prevented ∼175 million pneumococcal disease cases and >620,000 deaths in children <5 years of age.^2^ However, the invasive pneumococcal disease (IPD) burden remains substantial,^3^ with an estimated 40% of pediatric IPD cases caused by non-PCV13 serotypes.^4,5^

The 20-valent PCV (PCV20) contains conjugates for 7 additional serotypes beyond PCV13 (serotypes 8, 10A, 11A, 12F, 15B, 22F, and 33F); these additional serotypes were selected based on their prevalence in pediatric pneumococcal disease, serious and invasive disease potential, and antibiotic resistance.^4,6–13^ PCV20 is licensed for pneumococcal disease prevention in children from 6 weeks of age in many regions globally in a 4-dose (3 infant doses plus toddler dose [3+1]) or 3-dose series (2 infant doses plus toddler dose [2+1]).^14–18^ Licensure was based on results of 2 key phase 3 trials that investigated PCV20 safety and tolerability in infants and toddlers; a predominantly United States-based trial of a 3+1 schedule and a trial largely conducted in Europe of a 2+1 schedule.^19,20^ In both trials, PCV20 was well tolerated with a safety profile similar to that of PCV13 and elicited robust serotype-specific immune responses.^19,20^ For shared serotypes, however, immune responses after 2 primary PCV20 doses were somewhat lower compared with those after 2 PCV13 doses. In the 2+1 study, although noninferiority was met for IgG GMCs for 19 of the 20 serotypes 1 month after dose 3, importantly, after the infant series PCV20 missed noninferiority versus PCV13 for the IgG GMC coprimary endpoint for 4 of 13 matched serotypes.^20^ Although the clinical significance of missed noninferiority is unknown, further examination of the data is warranted to understand if PCV20 2-dose series immunogenicity can be improved by altering vaccination timing and to support the clinical effectiveness of the PCV20 2+1 schedule.^21,22^

The number and timing of primary infant PCV doses is important for development of robust early immune responses when infants are highly vulnerable to IPD.^23^ Studies on PCV13 have shown that 2 infant doses elicit lower immune responses post-primary series than 3 infant doses^24^ and that a 2-month interval between infant PCV doses elicits a stronger response and better protection than a shorter dosing interval.^25^ Additionally, administration of PCV13 primary doses at 3 and 5 months was reported to induce improved immune responses compared with administration at 2 and 4 months for 11 of 13 serotypes.^26^ However, immune responses to PCV13 after the toddler dose are generally similar for most serotypes regardless of number and timing of primary doses.^26^

In countries that recommend a 2+1 PCV schedule, infant doses may be given at 2 and 4 months (eg, Germany, France, Australia, Canada) or at 3 and 5 months (eg, the Netherlands).^27^ We therefore conducted a post hoc analysis to assess the impact of different infant dose timings on the immunogenicity, tolerability, and safety of PCV20 in healthy infants who received PCV20 or PCV13 in the key phase 3 trials.

## METHODS

### Studies and Participants

The study designs with inclusion and exclusion criteria of the phase 3, multicenter, double-blind, 3-dose (Study B7471012; NCT04546425) and 4-dose (Study B7471011; NCT04382326) trials have been reported previously.^19,20^ Briefly, Study B7471012 (2+1 study) included healthy infants from Europe and Australia who were 42−112 days of age at time of consent and born >36 weeks’ gestation. Participants were randomized 1:1 to receive 3 PCV20 or PCV13 doses: Dose 1 at enrollment, Dose 2 approximately 42−63 days after Dose 1, and Dose 3 (toddler dose) at 11−12 months of age. Study B7471011 (3+1 study) included healthy infants from the United States and Puerto Rico who were 42−98 days of age at time of consent and born >36 weeks’ gestation. Participants were randomized in a 1:1 ratio to receive 4 PCV20 or PCV13 doses at approximately 2, 4, 6, and 12−15 months of age. In both studies, participants’ parents/legal guardians provided written informed consent before enrollment.

### Assessments and Objectives

In the 2+1 study, immunogenicity endpoints were examined by timing of vaccination subgroup. The subgroup vaccinated at 2, 4, and 11−12 months (2,4m subgroup) included participants who received Doses 1 and 2 at 42–74 days of age and 85−134 days of age, respectively. The subgroup vaccinated at 3, 5, and 11−12 months (3,5m subgroup) included participants who received Doses 1 and 2 at 75−115 days of age and 135–180 days of age, respectively. Immune responses of the 2 subgroups were compared. Additionally, immune responses in the 2+1 study were compared with immune responses in the 3+1 study.

Immune responses for serotype-specific serum immunoglobulin G (IgG) concentrations were measured using a Luminex immunoassay.^28^ Immunogenicity endpoints included percentages of participants with predefined serotype-specific IgG concentrations 1 month after Dose 2 (2+1 study) and 1 month after Dose 3 (3+1 study). Predefined IgG concentrations were ≥0.35 μg/mL, except for serotypes 5 (≥0.23 μg/mL), 6B (≥0.10 μg/mL), and 19A (≥0.12 μg/mL), based on the Luminex assay bridging to the World Health Organization pneumococcal enzyme-linked immunosorbent assay (ELISA).^29^

Additionally, IgG geometric mean concentrations (GMCs) were compared by timing of vaccination post-primary series and post-toddler dose (1 month after Dose 2 and Dose 3 [2+1 study] and 1 month after Dose 3 and Dose 4 [3+1 study]). Lastly, in the 2+1 study subgroups, percentages of participants with predefined serotype-specific IgG concentrations and IgG GMCs were analyzed by receipt of maternal tetanus, diphtheria, and pertussis vaccine (Tdap). Post hoc safety endpoints by timing of vaccination subgroup for the 2+1 study were percentages of participants with local reactions (ie, redness, swelling, injection site pain) and systemic events (ie, fever, drowsiness, decreased appetite, irritability) reported within 7 days after each dose and adverse events (AEs) collected from Dose 1 through 1 month after Dose 2 and from Dose 3 through 1 month after Dose 3.

### Statistical Analyses

This was a descriptive post hoc analysis of 2 phase 3 clinical PCV infant studies. Exact 2-sided 95% CIs for percentages of participants achieving prespecified IgG concentrations were calculated based on the Clopper−Pearson method. Two-sided 95% CIs for the differences in percentages of participants achieving prespecified IgG concentrations were based on the Miettinen and Nurminen method. IgG GMCs and geometric mean ratios (GMRs) were calculated by exponentiating the mean logarithm of the concentrations or the ratios, respectively, with corresponding Student *t* distribution 95% CIs.

Safety data are presented descriptively as percentages for local reactions and systemic events. AEs are presented by system organ class as percentages with exact 2-sided Clopper−Pearson 95% CIs.

## RESULTS

### Participants

In the 2+1 study, 951/1207 (78.8%) randomized infants were included in this post hoc immunogenicity analysis. In the 2,4m subgroup, 330 infants received PCV20 and 326 infants received PCV13. In the 3,5m subgroup, 139 infants received PCV20 and 156 infants received PCV13. In the 3+1 study, 1636/1997 randomized infants (81.9%) were included in the immunogenicity evaluations (PCV20, n=833; PCV13, n=803).

In the 2+1 study sex, race, and ethnicity distributions were generally similar in the 2,4m and 3,5m subgroups (**Table 1**). In the 2,4m subgroup, median age at first vaccination was 55 days (range 43−74 days) in both the PCV20 and PCV13 groups. In the 3,5m subgroup, median age at first vaccination was 91 days (range 78−112 days) in the PCV20 group and 92 days (range 75−112 days) in the PCV13 group. In the 2,4m subgroup, nearly 75% of participants were enrolled in Poland (PCV20, 73.3%; PCV13, 73.0%). In the 3,5m subgroup, more than half of participants were enrolled in Finland (PCV20, 59.0%; PCV13, 55.8%) followed by Estonia (PCV20, 15.1%; PCV13, 14.7%). In the 2,4m subgroup, maternal Tdap was administered to 17.0% of mothers in the PCV20 group and 20.9% in the PCV13 group; corresponding percentages in the 3,5m subgroup were 5.0% and 7.1%, respectively. The 3+1 study demographic characteristics have been reported previously.^19^ Median age at Dose 1 was 64 days (range 42−79 days).^19^

**Table 1.**
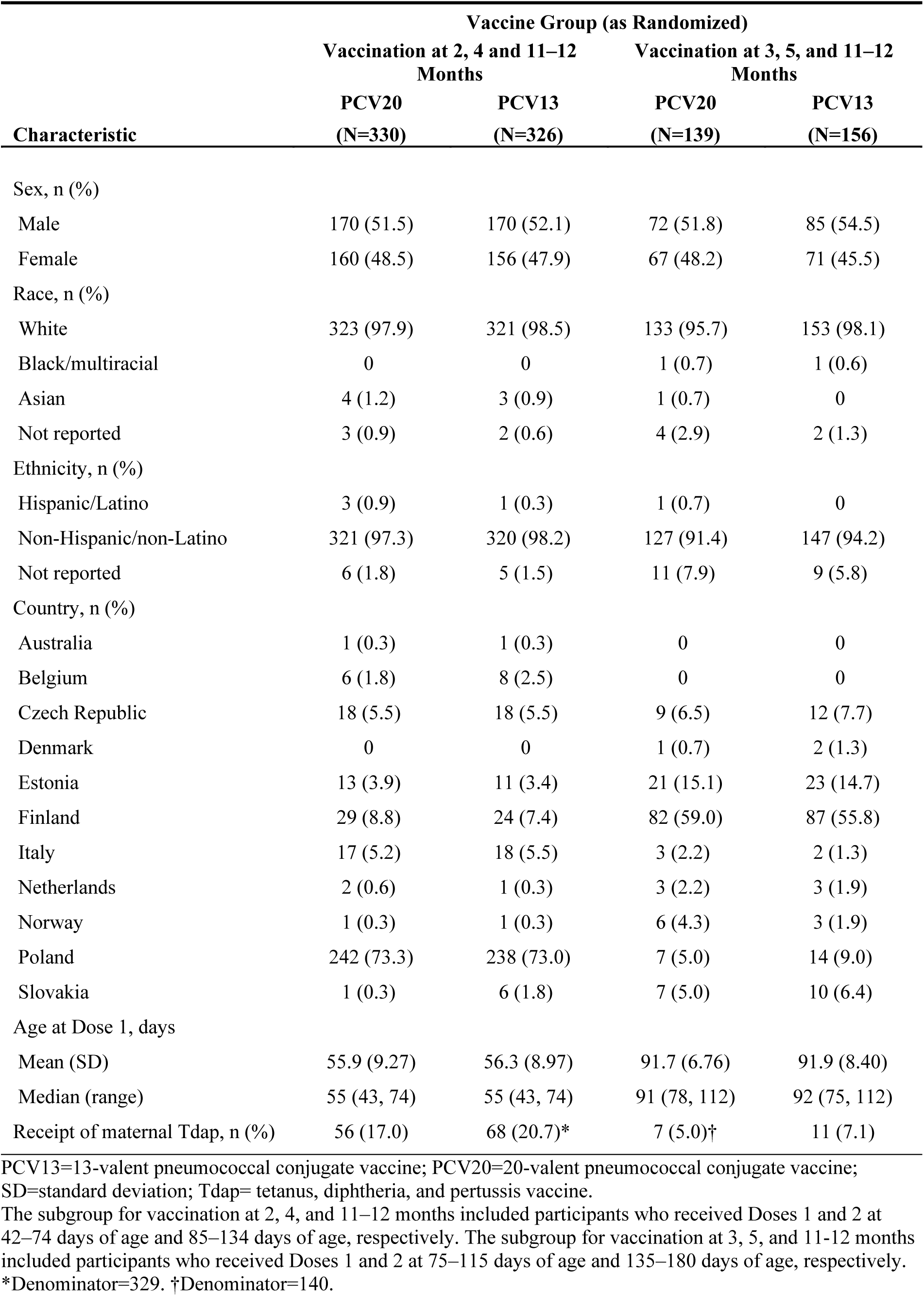
Demographic characteristics by timing of vaccination in the 2+1 study.

| Characteristic | Vaccine Group (as Randomized) |  |  |  |
| --- | --- | --- | --- | --- |
|  | Vaccination at 2, 4 and 11–12 Months |  | Vaccination at 3, 5, and 11–12 Months |  |
|  | PCV20<br>(N=330) | PCV13<br>(N=326) | PCV20<br>(N=139) | PCV13<br>(N=156) |
| Sex, n (%) |  |  |  |  |
| Male | 170 (51.5) | 170 (52.1) | 72 (51.8) | 85 (54.5) |
| Female | 160 (48.5) | 156 (47.9) | 67 (48.2) | 71 (45.5) |
| Race, n (%) |  |  |  |  |
| White | 323 (97.9) | 321 (98.5) | 133 (95.7) | 153 (98.1) |
| Black/multiracial | 0 | 0 | 1 (0.7) | 1 (0.6) |
| Asian | 4 (1.2) | 3 (0.9) | 1 (0.7) | 0 |
| Not reported | 3 (0.9) | 2 (0.6) | 4 (2.9) | 2 (1.3) |
| Ethnicity, n (%) |  |  |  |  |
| Hispanic/Latino | 3 (0.9) | 1 (0.3) | 1 (0.7) | 0 |
| Non-Hispanic/non-Latino | 321 (97.3) | 320 (98.2) | 127 (91.4) | 147 (94.2) |
| Not reported | 6 (1.8) | 5 (1.5) | 11 (7.9) | 9 (5.8) |
| Country, n (%) |  |  |  |  |
| Australia | 1 (0.3) | 1 (0.3) | 0 | 0 |
| Belgium | 6 (1.8) | 8 (2.5) | 0 | 0 |
| Czech Republic | 18 (5.5) | 18 (5.5) | 9 (6.5) | 12 (7.7) |
| Denmark | 0 | 0 | 1 (0.7) | 2 (1.3) |
| Estonia | 13 (3.9) | 11 (3.4) | 21 (15.1) | 23 (14.7) |
| Finland | 29 (8.8) | 24 (7.4) | 82 (59.0) | 87 (55.8) |
| Italy | 17 (5.2) | 18 (5.5) | 3 (2.2) | 2 (1.3) |
| Netherlands | 2 (0.6) | 1 (0.3) | 3 (2.2) | 3 (1.9) |
| Norway | 1 (0.3) | 1 (0.3) | 6 (4.3) | 3 (1.9) |
| Poland | 242 (73.3) | 238 (73.0) | 7 (5.0) | 14 (9.0) |
| Slovakia | 1 (0.3) | 6 (1.8) | 7 (5.0) | 10 (6.4) |
| Age at Dose 1, days |  |  |  |  |
| Mean (SD) | 55.9 (9.27) | 56.3 (8.97) | 91.7 (6.76) | 91.9 (8.40) |
| Median (range) | 55 (43, 74) | 55 (43, 74) | 91 (78, 112) | 92 (75, 112) |
| Receipt of maternal Tdap, n (%) | 56 (17.0) | 68 (20.7)* | 7 (5.0)† | 11 (7.1) |

PCV13=13-valent pneumococcal conjugate vaccine; PCV20=20-valent pneumococcal conjugate vaccine; SD=standard deviation; Tdap= tetanus, diphtheria, and pertussis vaccine.
The subgroup for vaccination at 2, 4, and 11–12 months included participants who received Doses 1 and 2 at 42–74 days of age and 85–134 days of age, respectively. The subgroup for vaccination at 3, 5, and 11–12 months included participants who received Doses 1 and 2 at 75–115 days of age and 135–180 days of age, respectively.
\*Denominator=329. †Denominator=140.

### Immunogenicity

#### Percentage of participants with predefined serotype-specific IgG concentrations

In the 2+1 study 1 month after Dose 2, the percentages of PCV20 recipients with predefined serotype-specific IgG concentrations in the PCV20 group were numerically higher for all 20 serotypes in the 3,5m subgroup compared with the 2,4m subgroup (**Figure 1**), with the lowest percentages for serotype 6B (20.4% in the 2,4m subgroup and 27.1% in the 3,5m subgroup) and the highest for serotype 8 (95.8% and 97.9%, respectively). The same pattern was observed in infants vaccinated with PCV13 for the 13 matched serotypes. For the 13 matched serotypes, the percentages of participants with predefined serotype-specific IgG concentrations 1 month after Dose 2 were higher in the PCV20 3,5m subgroup compared with the PCV13 2,4m subgroup except for serotypes 6B and 23F.

**Figure 1.**
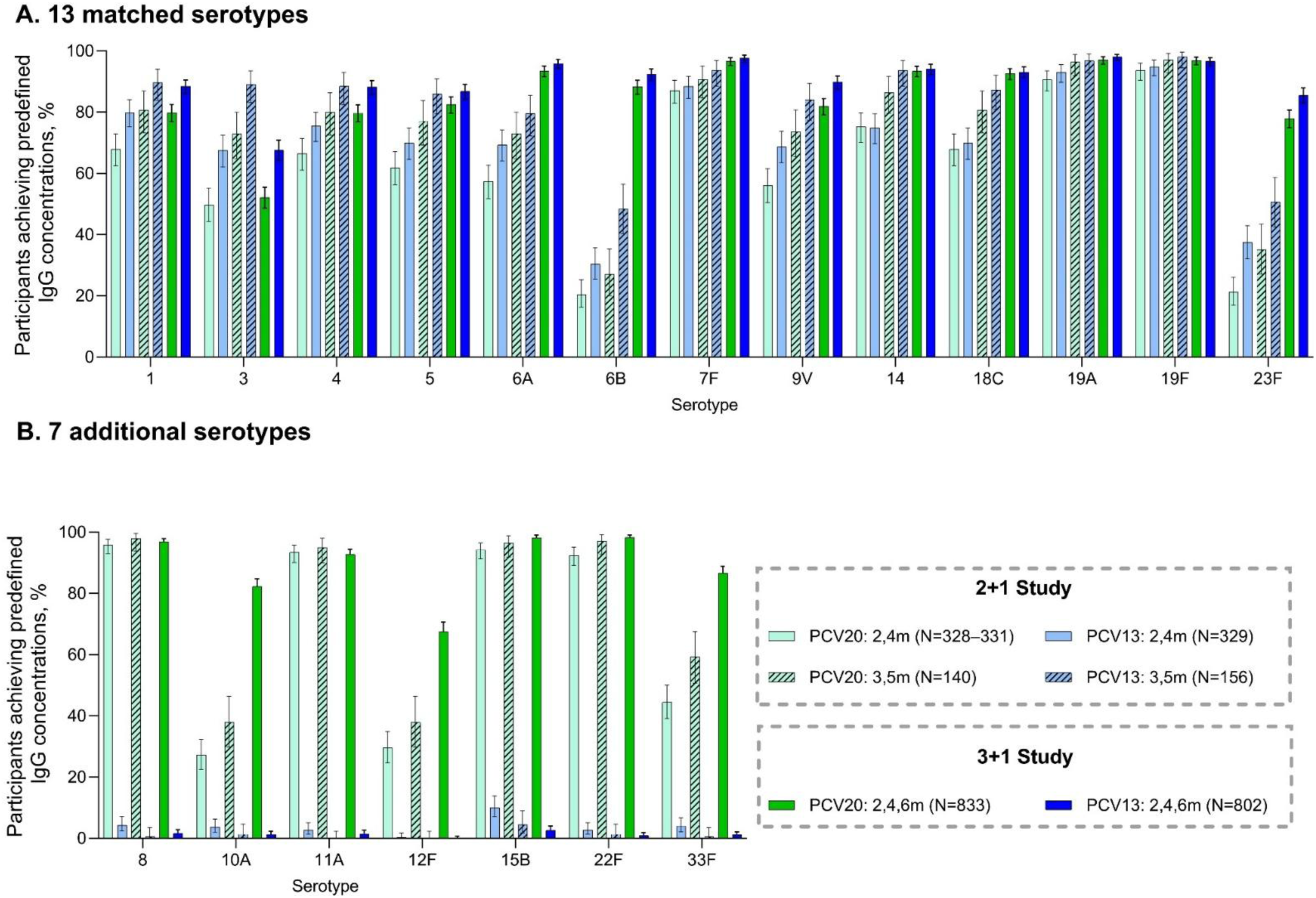
Percentages of participants with predefined serotype-specific IgG concentrations after the infant series by timing of vaccination 1 month after Dose 2 for the 2+1 study and 1 month after Dose 3 for the 3+1 study (A) for the 13 matched serotypes and (B) for the 7 additional serotypes Results are presented by primary series vaccination timing for the 2+1 study: the 2,4m subgroup included participants who received Doses 1 and 2 at 42–74 days of age and 85–134 days of age, respectively; the 3,5m subgroup included participants who received Doses 1 and 2 at 75–115 days of age and 135–180 days of age, respectively. In the 3+1 study, infant doses were given at 2, 4, and 6 months, respectively. Exact 2-sided CIs are based on the Clopper−Pearson method. IgG=immunoglobulin G; PCV13=13-valent pneumococcal conjugate vaccine; PCV20=20-valent pneumococcal conjugate vaccine.

The percentages of PCV20 recipients with predefined serotype-specific IgG concentrations 1 month after the third primary dose in the 3+1 study were substantially higher than after the second dose in the 2+1 study for serotypes 6A, 6B, and 23F among the 13 matched serotypes and serotypes 10A, 12F, and 33F among the 7 additional serotypes (**Figure 1**). For the other serotypes, the 3-dose priming series elicited similar percentages of responders compared with 2 priming doses in the 3,5m subgroup and generally higher percentages compared with the 2,4m subgroup.

The differences in percentages of participants with predefined serotype-specific IgG concentrations (PCV20 – PCV13) 1 month after Dose 2 in the 2+1 study are shown in **Figure 2A**; differences compared with PCV13 were generally similar between the 2,4m subgroup and the 3,5m subgroup. The corresponding differences in percentages of participants with predefined serotype-specific IgG concentrations (PCV20 minus PCV13) 1 month after Dose 3 in the 3+1 study are shown in **Figure 2B**.

**Figure 2.**
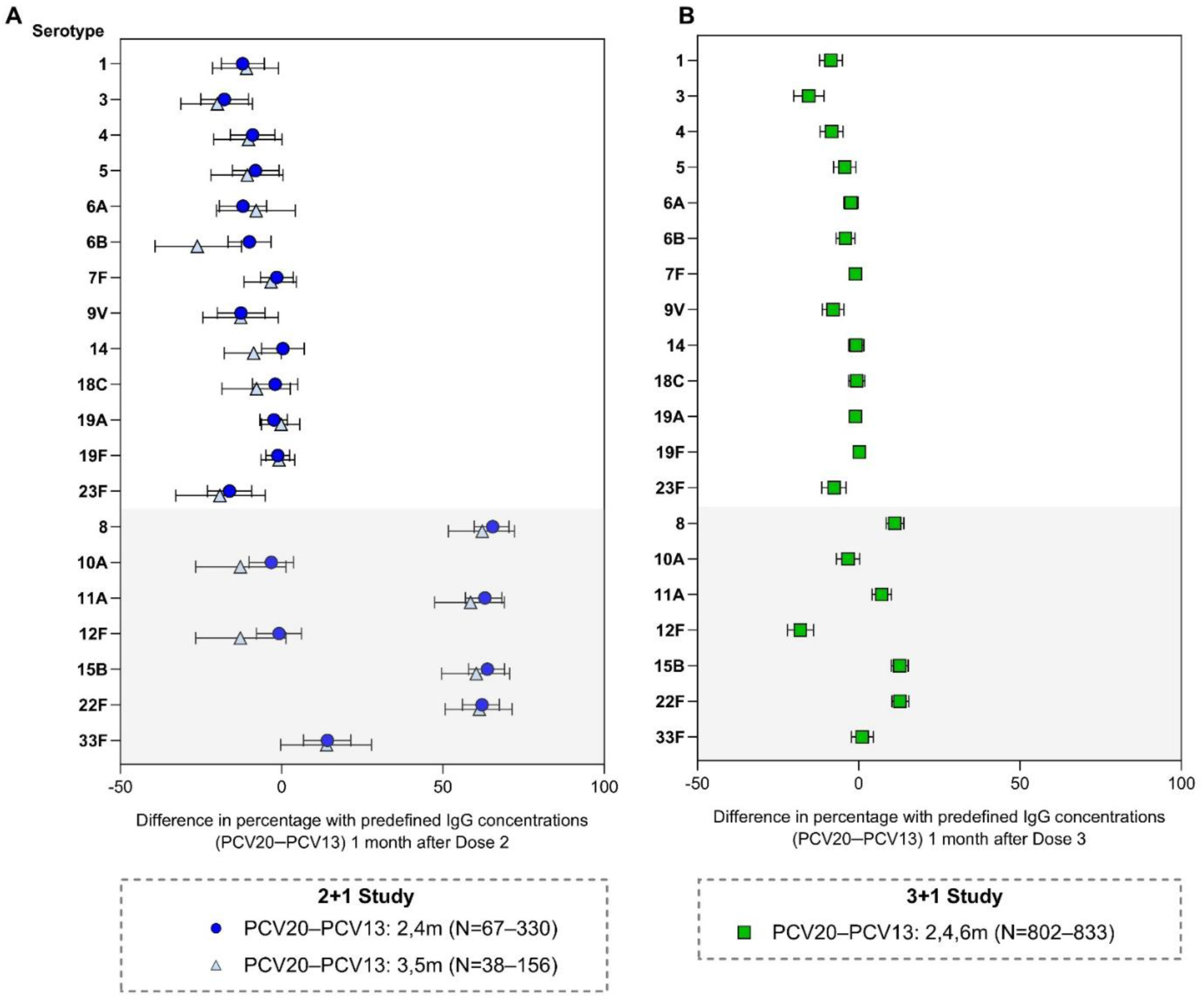
Differences (PCV20 − PCV13) with 2-sided 95% CIs in percentages of participants with predefined pneumococcal IgG levels (A) 1 month after Dose 2 in the 2+1 study and (B) 1 month after Dose 3 in the 3+1 study. The predefined pneumococcal IgG level for all serotypes is ≥0.35 μg/mL except for the following: 5 (≥0.23 μg/mL), 6B (≥0.10 μg/mL), and 19A (≥0.12 μg/mL). For the PCV13 matched serotypes, the compared results are from the corresponding serotype in the PCV13 group. For the 7 additional serotypes (gray shading), the compared results are from the PCV13 serotype with the lowest percentage in the PCV13 group not including serotype 3 (serotype 6B in panel A and serotype 23F in panel B). Results are presented by primary series vaccination timing for the 2+1 study: the 2,4m subgroup included participants who received Doses 1 and 2 at 42–74 days of age and 85–134 days of age, respectively; the 3,5m subgroup included participants who received Doses 1 and 2 at 75–115 days of age and 135–180 days of age, respectively. IgG=immunoglobulin G; N, the range of participants across PCV20 and PCV13 groups with an IgG concentration ≥ the predefined level for the given serotype. PCV13=13-valent pneumococcal conjugate vaccine; PCV20=20-valent pneumococcal conjugate vaccine.

### IgG GMCs

One month after PCV20 Dose 2 in the 2+1 study, IgG GMCs were numerically higher in the 3,5m subgroup compared with the 2,4m subgroup for all serotypes (**Figure 3**), with the lowest IgG GMCs observed for serotype 6B (0.03 µg/mL in the 2,4m subgroup and 0.05 µg/mL in the 3,5m subgroup) and the highest for serotype 15B (3.0 µg/mL in the 2,4m subgroup and 4.6 µg/mL in the 3,5m subgroup). The same pattern was observed in infants vaccinated with PCV13 for the 13 matched serotypes. For the 13 matched serotypes, observed IgG GMCs were generally similar in the PCV20 3,5m subgroup and the PCV13 2,4m subgroup, and higher with PCV20 3,5m for serotypes 3, 5, 14, and 18C.

**Figure 3.**
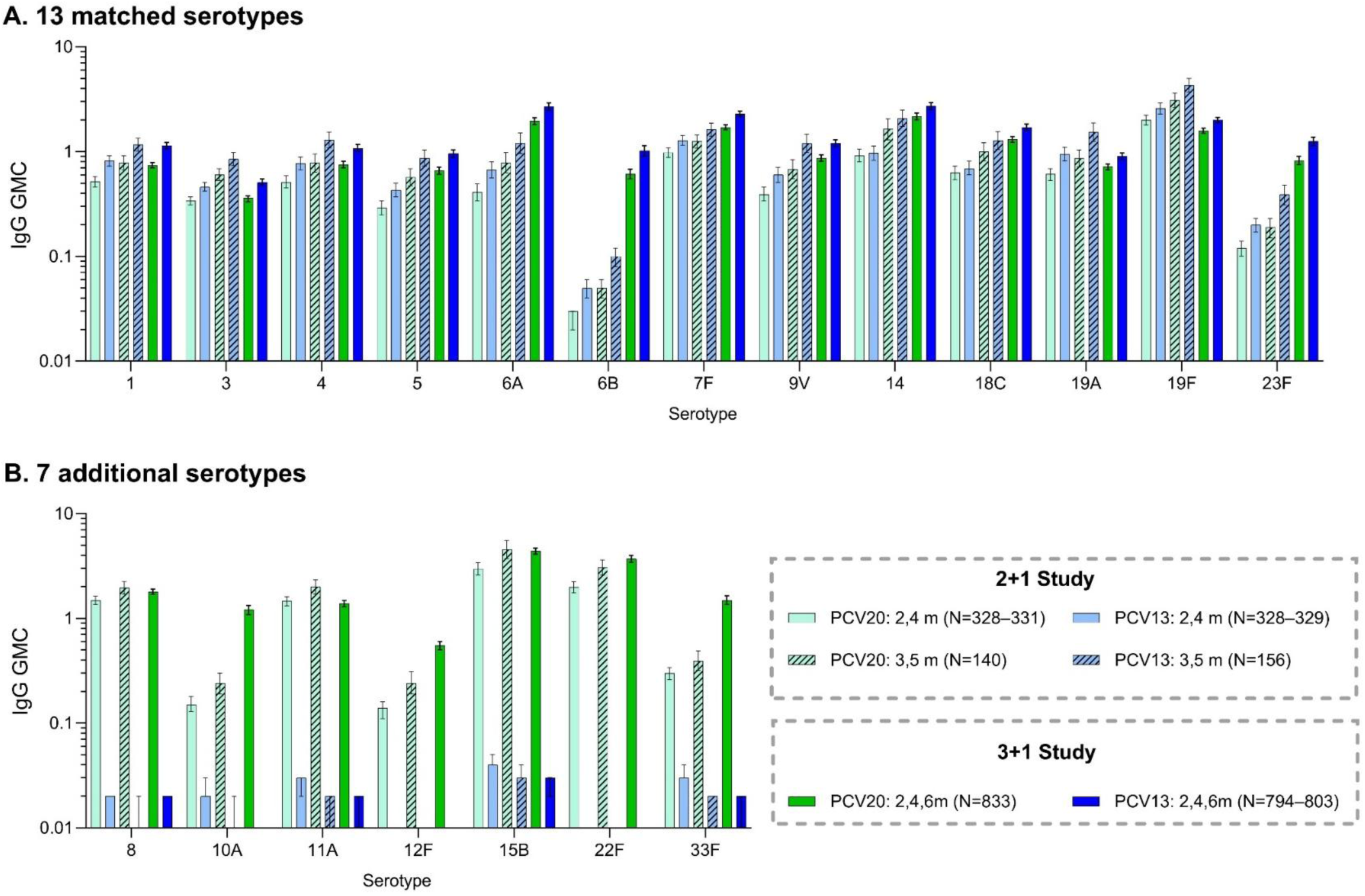
IgG GMCs (2-sided 95% CIs) after the infant series by timing of vaccination 1 month after Dose 2 for the 2+1 study and 1 month after Dose 3 for the 3+1 study (A) for the 13 matched serotypes and (B) for the 7 additional serotypes. Results are presented by primary series vaccination timing for the 2+1 study: the 2,4m subgroup included participants who received Doses 1 and 2 at 42–74 days of age and 85–134 days of age, respectively; the 3,5m subgroup included participants who received Doses 1 and 2 at 75–115 days of age and 135–180 days of age, respectively. In the 3+1 study, doses were given at 2, 4, and 6 months, respectively. GMCs and 2-sided CIs were calculated by exponentiating the mean logarithm of the concentrations and the corresponding CIs (based on the Student *t* distribution). GMC=geometric mean concentrations; IgG=immunoglobulin G; PCV13=13-valent pneumococcal conjugate vaccine; PCV20=20-valent pneumococcal conjugate vaccine.

Observed IgG GMCs after PCV20 Dose 3 in the 3+1 study were markedly higher than after Dose 2 in the 2+1 study for serotypes 6B and 23F from the 13 matched serotypes, and 10A, 12F, and 33F for the 7 additional serotypes (**Figure 3**). For the other serotypes, IgG GMCs after a 3-dose primary series were generally similar to those in the 3,5m subgroup and higher than those in the 2,4 m subgroup.

One month after the toddler dose, PCV20 and PCV13 elicited strong immune responses with serotype-specific IgG GMCs being generally similar in the 2+1 (both 2,4m and 3,5m subgroups) and 3+1 schedules (**Figure 4**). IgG GMCs were numerically higher in the PCV13 2,4m subgroup than in the PCV20 3,5m subgroup for all 13 matched serotypes. The IgG GMRs (PCV20/PCV13) 1 month after Dose 2 and 1 month after Dose 3 in the 2+1 study were generally similar between the 2,4m and 3,5m subgroups (**Supplemental Content 1**). Corresponding IgG GMRs 1 month after Dose 3 and 1 month after Dose 4 in the 3+1 study are shown in **Supplemental Content 2**.

**Figure 4.**
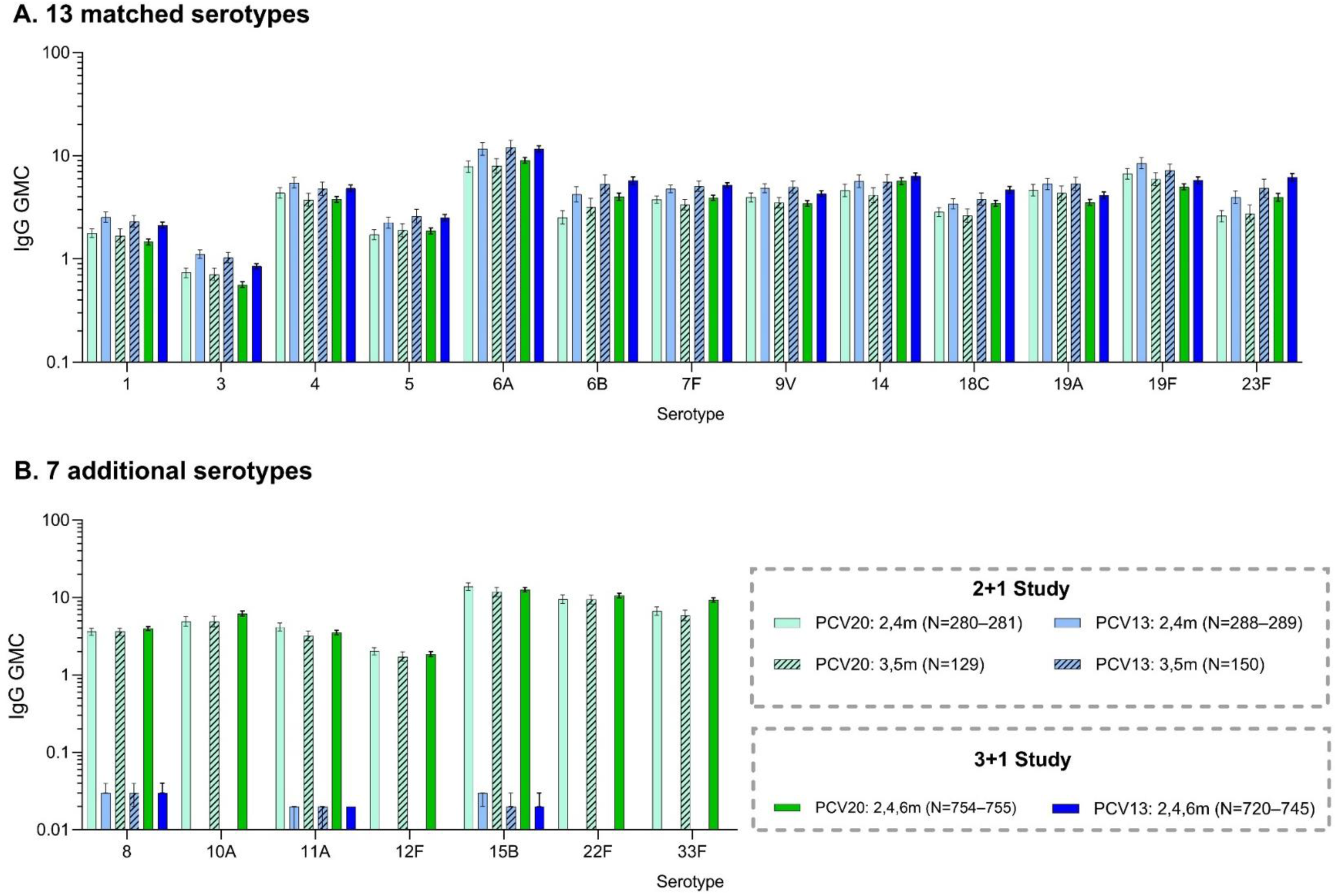
IgG GMCs (2-sided 95% CIs) after the toddler dose by timing of vaccination 1 month after Dose 3 for the 2+1 study and 1 month after Dose 4 for the 3+1 study (A) for the 13 matched serotypes and (B) for the 7 additional serotypes Results are presented by primary series vaccination timing for the 2+1 study: the 2,4m subgroup included participants who received Doses 1, 2, and 3 at 42–74 days of age, 85–134 days of age, and 11–12 months, respectively; the 3,5m subgroup included participants who received Doses 1, 2, and 3 at 75–115 days of age, 135–180 days of age, and 11–12 months, respectively. In the 3+1 study, doses were given at 2, 4, 6, and 12–15 months, respectively. GMCs and 2-sided CIs were calculated by exponentiating the mean logarithm of the concentrations and the corresponding CIs (based on the Student *t* distribution). GMC=geometric mean concentrations; IgG=immunoglobulin G; PCV13=13-valent pneumococcal conjugate vaccine; PCV20=20-valent pneumococcal conjugate vaccine.

### Maternal Tdap vaccination

In the 2+1 study 2,4m subgroup, maternal Tdap vaccination was associated with generally lower IgG responses compared with no maternal Tdap for all serotypes in PCV20- and PCV13-vaccinated participants (**Supplemental Content 3**). This trend was not observed in the 3,5m subgroup, although the small numbers of mothers vaccinated with Tdap in the 3,5m subgroups preclude comparisons.

### Safety

In the 2+1 study post hoc safety analysis, local reactions and systemic events were generally reported at similar rates across groups with no obvious or consistent differences observed with the 2,4m subgroups compared with the 3,5m subgroups (**Supplemental Content 4**).

Similarly, AEs were reported at generally similar frequencies across groups, with AEs most commonly reported in the infections and infestations system organ class (**Supplemental Content 5**). Safety data for the 3+1 study have been reported previously.^19^ Briefly, a PCV20 3+1 regimen was well tolerated with a safety profile similar to that of PCV13.

## DISCUSSION

This post hoc analysis from 2 phase 3 PCV20 immunogenicity trials compared the immunogenicity of 3 pediatric primary immunization schedules: a 2+1 schedule with primary doses administered either at 2 and 4 months, or 3 and 5 months, and a 3+1 schedule with primary doses administered at 2, 4, and 6 months. Immune responses after the toddler doses, were robust and generally comparable in all 3 schedules, providing confidence in consistent population protection across schedules, given the importance of the toddler doses in inducing memory responses^30^ and reducing colonization.^31,32^

Immunogenicity after the infant series varied by schedule. Immune responses were higher after 2 PCV20 infant doses given at 3 and 5 months than at 2 and 4 months, and the same was observed for PCV13. Our results are consistent with a randomized controlled trial comparing immune responses of 4 different dosing regimens of PCV13 (infant doses at 2, and 4 months; 3, and 5 months; 2, 4, and 6 months; or 2, 3, and 4 months) in which a 3- and 5-month infant schedule was superior to a 2- and 4-month schedule for 11 of 13 serotypes.^26^

Importantly, post-primary responses to PCV20 administered at 3 and 5 months were similar to responses to PCV13 administered at 2 and 4 months for most of the 13 matched serotypes, and higher for serotypes 3, 5, 14, and 18C. Although this comparison was descriptive, with 3-and 5-month and 2- and 4-month schedules administered in different populations based on the national immunization schedule at each trial site, these data suggest that a 1-month delay in initiating the PCV20 primary series may provide similar direct protection during infancy as PCV13 for the matched serotypes, while expanding coverage to 7 additional highly prevalent and medically important serotypes. These results may help address concerns regarding lower immunogenicity after the primary series of a 2+1 schedule of PCV20.^22^

The 3+1 schedule with primary doses administered at 2, 4, and 6 months generally induced the highest IgG responses after the infant series compared with both 2-dose primary schedules, especially for serotypes 6A, 6B, 23F, 10A, 12F, and 33F. However, for the other serotypes, responses were generally similar after infant doses in the 3,5m 2+1 subgroup and the 2,4,6m 3+1 schedule. In our analysis, PCV20 elicited strong toddler dose immune responses at 11–12 months of age in both 2-dose primary regimens in the 2+1 study, leading to similar IgG GMCs to those after the toddler dose in the 3+1 study for all serotypes, including for serotypes 6A, 6B, 23F, 10A, 12F, and 33F. These results are consistent with the randomized controlled trial of 4 different dosing schedules of PCV13 in which there were no statistically significant differences in antibody levels between schedules after the booster dose for most serotypes.^26^

Although a 3+1 PCV schedule induces more robust immune responses than a 2+1 schedule against several serotypes and is associated with moderately higher direct protection during the first year of life,^33,34^ several studies have shown that direct protection for the complete series, as well as population-level protection, are similar for both schedules.^33,35^ Hence, there may be little increased benefit of a 3+1 schedule in terms of pneumococcal disease reduction in populations with high booster uptake once herd effects are established.^36,37^

In our analysis, maternal Tdap vaccination was associated with generally lower IgG responses compared with no maternal Tdap for all serotypes in the 2+1 study 2,4m subgroups; in the 3,5m subgroups, the number of maternal Tdap recipients was too small to make any meaningful comparisons. Although we cannot exclude that maternal Tdap may have contributed to the 3,5m vs. 2,4m comparison findings, our results are consistent with the PCV13 dosing schedule trial,^26^ suggesting negligible bias. Maternal Tdap is recommended in several European countries (**Supplemental Content 6**), and may blunt infant PCV responses, although reduced immunogenicity may not be clinically significant.^38^

In this post hoc analysis from the 2+1 study, vaccination timing did not affect PCV20 tolerability or safety, with local reactions, systemic events, and AEs being generally similar across the 2,4m and 3,5m subgroups, similar to those of PCV13 and consistent with the primary safety analyses from both the 2+1 and 3+1 studies.^19,20^

A limitation of this descriptive post hoc analysis is that it was not powered to detect differences between timing of vaccination subgroups or between studies. Additionally, the subgroup analysis for the 2 different administration regimens of the 2-dose infant series was not protected by randomization and we cannot exclude residual confounding.

In conclusion, IgG responses after 2 PCV20 doses given at 3 and 5 months were similar to those after administration of PCV13 at 2 and 4 months for the matched serotypes. Responses were also similar to those after 3 PCV20 doses at 2,4 and 6 months for 14/20 serotypes; the 3-dose primary schedule was advantageous for serotypes 6A, 6B, 23F, 10A, 12F and 33F. Practical considerations, including costs and crowding of the infant immunization schedule, have made 2+1 schedules the preferred choice in many regions and countries.^35,39^ Our analysis provides evidence to countries with a 2-dose primary schedule wishing to optimize coverage, suggesting that a 1-month delay in PCV20 dosing may provide more robust direct protection against vaccine-type pneumococcal disease during late infancy. Regardless of primary schedule, increasing PCV20 uptake and booster dose compliance is critical to achieve the full public health benefits of the infant vaccination program, including both direct and indirect protection against invasive pneumococcal disease, pneumonia, and acute otitis media.

## Data Availability

Upon request, and subject to review, Pfizer will provide the data that support the findings of these studies. Subject to certain criteria, conditions, and exceptions, Pfizer may also provide access to the related individual de-identified participant data. See https://www.pfizer.com/science/clinical-trials/trial-data-and-results for more information.

## Acknowledgments

The authors thank the children and their caregivers who participated in these studies and the investigators and study site staff. Medical writing support was provided by Sheena Hunt, PhD, of ICON (Blue Bell, PA) and was funded by Pfizer Inc.

## Conflicts of Interest and Source of Funding

All authors are Pfizer employees and may hold stock or stock options. This study was sponsored by Pfizer Inc.

## Notes

### Clinical Trial

NCT04546425
NCT04382326

### Author Declarations

Ethics committee approvals for B7471011 study (Institutional Review Board or Ethics Committee, Country): San Juan Hospital Institutional Review Board, Puerto Rico; Advarra, USA; Western Institutional Review Board, USA; Institutional Review Boards University of Louisville, USA; East Carolina University, University & Medical Center, USA; Creighton University, USA; Kaiser Permanente Northern California, USA; University of Tennessee Health Science Center, USA.

