## Appendix for "Safety and Immunogenicity of 20-Valent PCV According to Number and Timing of Primary Series Doses": Kline-medRxiv Appendix.pdf

### Supplementary Appendix

### Supplemental Content 1. IgG GMRs (PCV20/PCV13) and 2-sided 95% CIs (A) 1 month after Dose 2 and (B) 1 month after Dose 3 in the 2+1 study

For the PCV13 matched serotypes, the compared results are from the corresponding serotype in the PCV13 group. For the 7 additional serotypes (gray shading), the compared results are from the PCV13 serotype with the lowest GMC, not including serotype 3 in the PCV13 group (serotype 6B in both subgroups in panel A; serotype 5 in the 2,4m subgroup and serotype 1 in the 3,5m subgroup in panel B). Results are presented by primary series vaccination timing for the 2+1 study: the 2,4m subgroup included participants who received Doses 1 and 2 at 42–74 days of age and 85–134 days of age, respectively; the 3,5m subgroup included participants who received Doses 1 and 2 at 75–115 days of age and 135–180 days of age, respectively. GMC=geometric mean concentration; GMR=geometric mean ratio; IgG=immunoglobulin G; PCV13=13-valent pneumococcal conjugate vaccine; PCV20=20-valent pneumococcal conjugate vaccine.

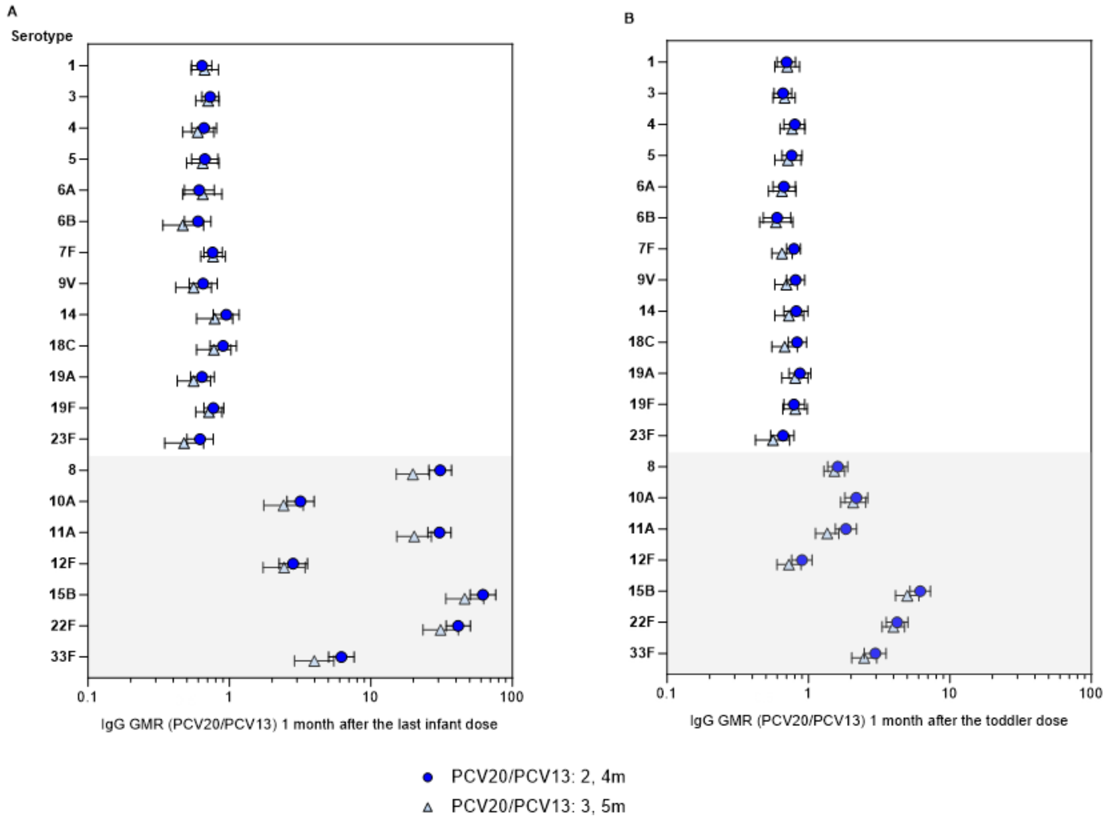

### Supplemental Content 2. IgG GMRs (PCV20/PCV13) and 2-sided 95% CIs (A) 1 month after Dose 3 and (B) 1 month Dose 4 in the 3+1 study

These data have been presented previously in a different format.[1] For the PCV13 matched serotypes, the compared results are from the corresponding serotype in the PCV13 group. For the 7 additional serotypes (grey shading), the compared results are from the PCV13 serotype with the lowest GMC, not including serotype 3 in the PCV13 group (serotype 19A in panel A; serotype 1 in panel B). GMC=geometric mean concentration; GMR=geometric mean ratio; IgG=immunoglobulin G; PCV13=13-valent pneumococcal conjugate vaccine; PCV20=20-valent pneumococcal conjugate vaccine.

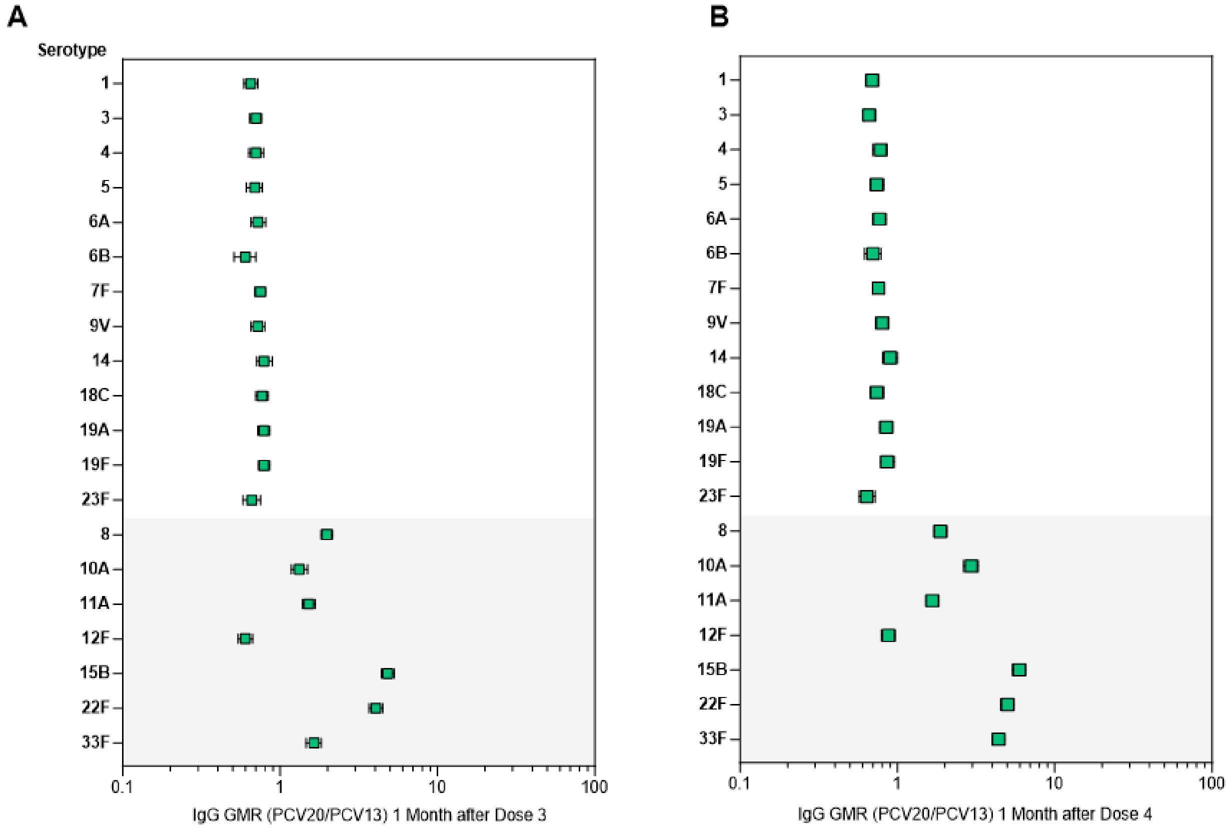

[1] Senders S, et al. A phase three study of the safety and immunogenicity of a four-dose series of 20-valent pneumococcal conjugate vaccine in healthy infants. *Pediatr Infect Dis J*. 2024;43(6):596-603. doi: 10.1097/INF.0000000000004334.

### Supplemental Content 3. Percentage of participants with predefined pneumococcal IgG concentrations for vaccine serotypes by vaccination timing and maternal Tdap use 1 month after Dose 2 for the 2+1 study

| | | Vaccine group (as randomized) | | | | | | | |
|---|---|---|---|---|---|---|---|---|---|
| | | Vaccination at 2, 4, and 11-12 months | | | | Vaccination at 3, 5, and 11-12 months | | | |
| | | With Tdap | | Without Tdap | | With Tdap | | Without Tdap | |
| Serotype | Predefined Level (µg/mL) | PCV20 (N=56) % (95% CI) | PCV13 (N=68) % (95% CI) | PCV20 (N=274) % (95% CI) | PCV13 (N=261) % (95% CI) | PCV20 (N=7) % (95% CI) | PCV13 (N=11) % (95% CI) | PCV20 (N=133) % (95% CI) | PCV13 (N=145) % (95% CI) |
| **Matched serotypes** | | | | | | | | | |
| 1 | ≥0.35 | 42.9 (29.7, 56.8) | 61.8 (49.2, 73.3) | 73.0 (67.3, 78.2) | 84.7 (79.7, 88.8) | 100.0 (59.0, 100.0) | 81.8 (48.2, 97.7) | 79.7 (71.9, 86.2) | 90.3 (84.3, 94.6) |
| 3 | ≥0.35 | 26.8 (15.8, 40.3) | 54.4 (41.9, 66.5) | 54.4 (48.3, 60.4) | 70.9 (65.0, 76.3) | 71.4 (29.0, 96.3) | 63.6 (30.8, 89.1) | 72.9 (64.5, 80.3) | 91.0 (85.2, 95.1) |
| 4 | ≥0.35 | 33.9 (21.8, 47.8) | 57.4 (44.8, 69.3) | 73.0 (67.3, 78.2) | 80.1 (74.7, 84.7) | 85.7 (42.1, 99.6) | 81.8 (48.2, 97.7) | 79.7 (71.9, 86.2) | 89.0 (82.7, 93.6) |
| 5 | ≥0.23 | 48.2 (34.7, 62.0) | 50.0 (37.6, 62.4) | 64.6 (58.6, 70.3) | 75.1 (69.4, 80.2) | 71.4 (29.0, 96.3) | 54.5 (23.4, 83.3) | 77.4 (69.4, 84.2) | 88.3 (81.9, 93.0) |
| 6A | ≥0.35 | 42.9 (29.7, 56.8) | 61.8 (49.2, 73.3) | 60.2 (54.2, 66.1) | 71.3 (65.4, 76.7) | 85.7 (42.1, 99.6) | 54.5 (23.4, 83.3) | 72.2 (63.7, 79.6) | 81.4 (74.1, 87.4) |
| 6B | ≥0.10 | 8.9 (3.0, 19.6) | 17.6 (9.5, 28.8) | 22.8 (17.9, 28.2) | 33.7 (28.0, 39.8) | 42.9 (9.9, 81.6) | 18.2 (2.3, 51.8) | 26.3 (19.1, 34.7) | 50.7 (42.2, 59.1) |
| 7F | ≥0.35 | 82.1 (69.6, 91.1) | 77.9 (66.2, 87.1) | 88.0 (83.5, 91.6) | 91.2 (87.1, 94.3) | 100.0 (59.0, 100.0) | 90.9 (58.7, 99.8) | 90.2 (83.9, 94.7) | 93.8 (88.5, 97.1) |
| 9V | ≥0.35 | 37.5 (24.9, 51.5) | 52.9 (40.4, 65.2) | 59.9 (53.8, 65.7) | 72.8 (67.0, 78.1) | 57.1 (18.4, 90.1) | 63.6 (30.8, 89.1) | 74.4 (66.2, 81.6) | 85.5 (78.7, 90.8) |
| 14 | ≥0.35 | 64.3 (50.4, 76.6) | 63.2 (50.7, 74.6) | 77.4 (72.0, 82.2) | 77.8 (72.2, 82.7) | 85.7 (42.1, 99.6) | 90.9 (58.7, 99.8) | 86.5 (79.5, 91.8) | 93.8 (88.5, 97.1) |
| 18C | ≥0.35 | 46.4 (33.0, 60.3) | 58.8 (46.2, 70.6) | 72.3 (66.6, 77.5) | 72.8 (67.0, 78.1) | 85.7 (42.1, 99.6) | 72.7 (39.0, 94.0) | 80.5 (72.7, 86.8) | 88.3 (81.9, 93.0) |
| 19A | ≥0.12 | 87.5 (75.9, 94.8) | 91.2 (81.8, 96.7) | 91.2 (87.2, 94.3) | 93.5 (89.8, 96.2) | 85.7 (42.1, 99.6) | 90.9 (58.7, 99.8) | 97.0 (92.5, 99.2) | 97.2 (93.1, 99.2) |
| 19F | ≥0.35 | 87.5 (75.9, 94.8) | 88.2 (78.1, 94.8) | 94.9 (91.6, 97.2) | 96.6 (93.6, 98.4) | 100.0 (59.0, 100.0) | 90.9 (58.7, 99.8) | 97.0 (92.5, 99.2) | 98.6 (95.1, 99.8) |
| 23F | ≥0.35 | 12.5 (5.2, 24.1) | 26.5 (16.5, 38.6) | 23.0 (18.1, 28.4) | 40.2 (34.2, 46.5) | 14.3 (0.4, 57.9) | 45.5 (16.7, 76.6) | 36.1 (27.9, 44.9) | 51.0 (42.6, 59.4) |
| **7 Additional serotypes** | | | | | | | | | |
| 8 | ≥0.35 | 91.1 (80.4, 97.0) | 0 (0.0, 5.4) | 96.7 (93.9, 98.5) | 5.4 (3.0, 8.8) | 100.0 (59.0, 100.0) | 0 (0.0, 28.5) | 97.7 (93.5, 99.5) | 0.7 (0.0, 3.8) |
| 10A | ≥0.35 | 16.1 (7.6, 28.3) | 8.8 (3.3, 18.2) | 29.5 (24.1, 35.2) | 2.3 (0.8, 4.9) | 14.3 (0.4, 57.9) | 0 (0.0, 28.5) | 39.1 (30.8, 47.9) | 1.4 (0.2, 4.9) |
| 11A | ≥0.35 | 85.7 (73.8, 93.6) | 0 (0.0, 5.3) | 94.9 (91.6, 97.2) | 3.4 (1.6, 6.4) | 100.0 (59.0, 100.0) | 0 (0.0, 28.5) | 94.7 (89.5, 97.9) | 0 (0.0, 2.5) |
| 12F | ≥0.35 | 14.3 (6.4, 26.2) | 0 (0.0, 5.3) | 32.7 (27.2, 38.6) | 0.4 (0.0, 2.1) | 42.9 (9.9, 81.6) | 0 (0.0, 28.5) | 37.6 (29.3, 46.4) | 0 (0.0, 2.5) |
| 15B | ≥0.35 | 87.5 (75.9, 94.8) | 7.4 (2.4, 16.3) | 95.6 (92.5, 97.7) | 10.7 (7.2, 15.1) | 100.0 (59.0, 100.0) | 0 (0.0, 28.5) | 96.2 (91.4, 98.8) | 4.8 (2.0, 9.7) |
| 22F | ≥0.35 | 85.7 (73.8, 93.6) | 1.5 (0.0, 7.9) | 93.8 (90.3, 96.4) | 3.1 (1.3, 5.9) | 100.0 (59.0, 100.0) | 0 (0.0, 28.5) | 97.0 (92.5, 99.2) | 1.4 (0.2, 4.9) |
| 33F | ≥0.35 | 16.1 (7.6, 28.3) | 2.9 (0.4, 10.2) | 50.4 (44.3, 56.4) | 4.2 (2.1, 7.4) | 57.1 (18.4, 90.1) | 0 (0.0, 28.5) | 59.4 (50.5, 67.8) | 0.7 (0.0, 3.8) |

IgG=immunoglobulin G; PCV13=13-valent pneumococcal conjugate vaccine; PCV20=20-valent pneumococcal conjugate vaccine; SD, standard deviation; Tdap, tetanus, diphtheria, and pertussis vaccine.

The subgroup for vaccination at 2, 4, and 11–12 months included participants who received Doses 1 and 2 at 42–74 days of age and 85–134 days of age, respectively. The subgroup for vaccination at 3, 5, and 11–12 months included participants who received Doses 1 and 2 at 75–115 days of age and 135–180 days of age, respectively. Exact 2-sided CI, based on the Clopper and Pearson method.

### Supplemental Content 4. Percentages of participants with reported (A) local reactions and (B) systemic events within 7 days after each dose by timing of vaccination subgroup for the 2+1 study

Results are for the safety population in the 2+1 study. Results are presented by primary series vaccination timing: the 2,4m subgroup included participants who received Doses 1, 2, and 3 at 42–74 days of age, 85–134 days of age, and 11–12 months, respectively; the 3,5m subgroup included participants who received Doses 1, 2, and 3 at 75–115 days of age, 135–180 days of age, and 11–12 months, respectively. D=dose; PCV13=13-valent pneumococcal conjugate vaccine; PCV20=20-valent pneumococcal conjugate vaccine.

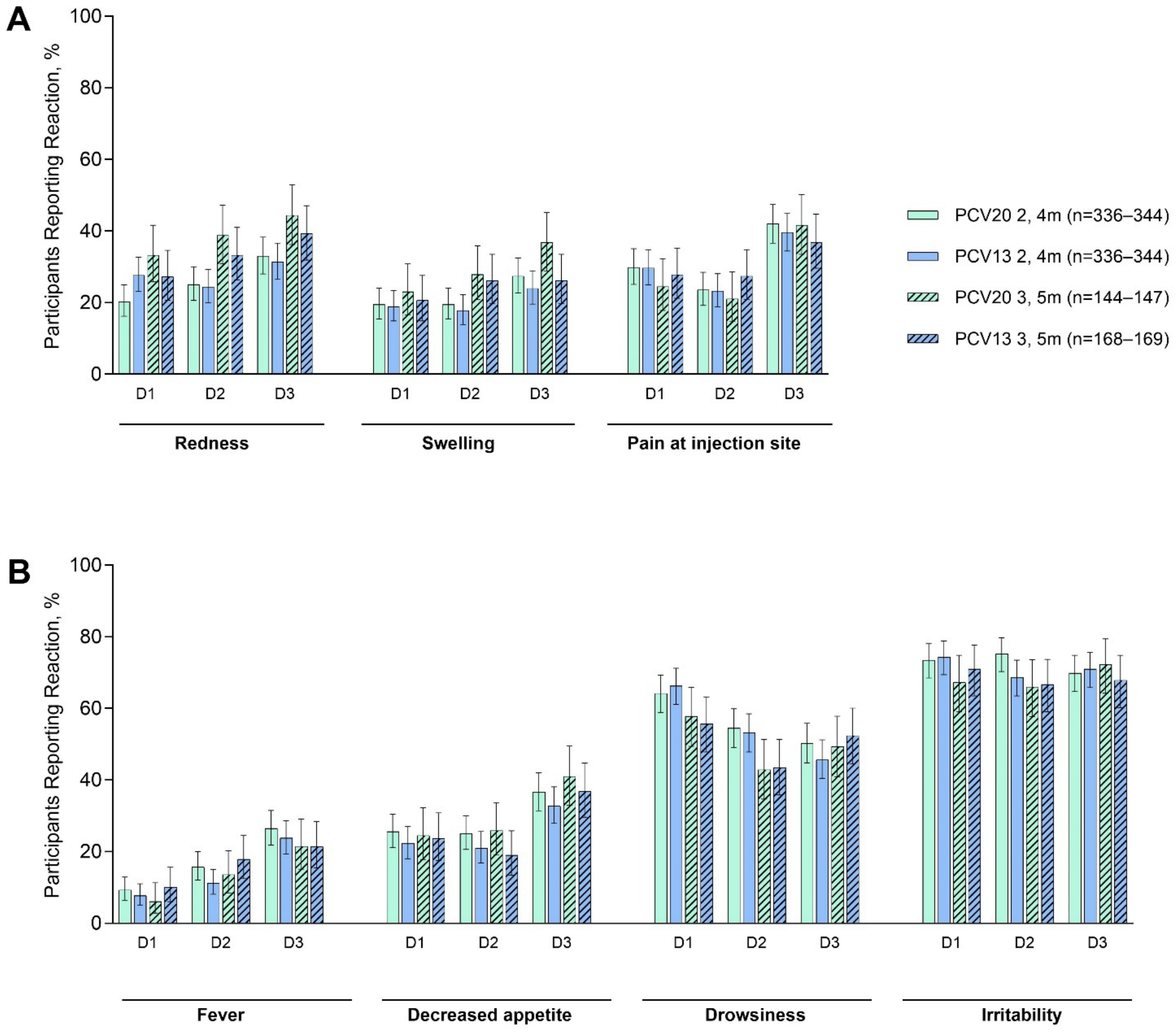

### Supplemental Content 5. Percentages of participants with AEs overall and by system organ class from the 2+1 study (A) from Dose 1 to 1 month after Dose 2 and (B) from Dose 3 to 1 month after Dose 3

Results are presented by primary series vaccination timing for the 2+1 study: the 2,4m subgroup included participants who received Doses 1, 2, and 3 at 42–74 days of age, 85–134 days of age, and 11–12 months, respectively; the 3,5m subgroup included participants who received Doses 1, 2, and 3 at 75–115 days of age, 135–180 days of age, and 11–12 months, respectively. AE=adverse event; PCV13=13-valent pneumococcal conjugate vaccine; PCV20=20-valent pneumococcal conjugate vaccine.

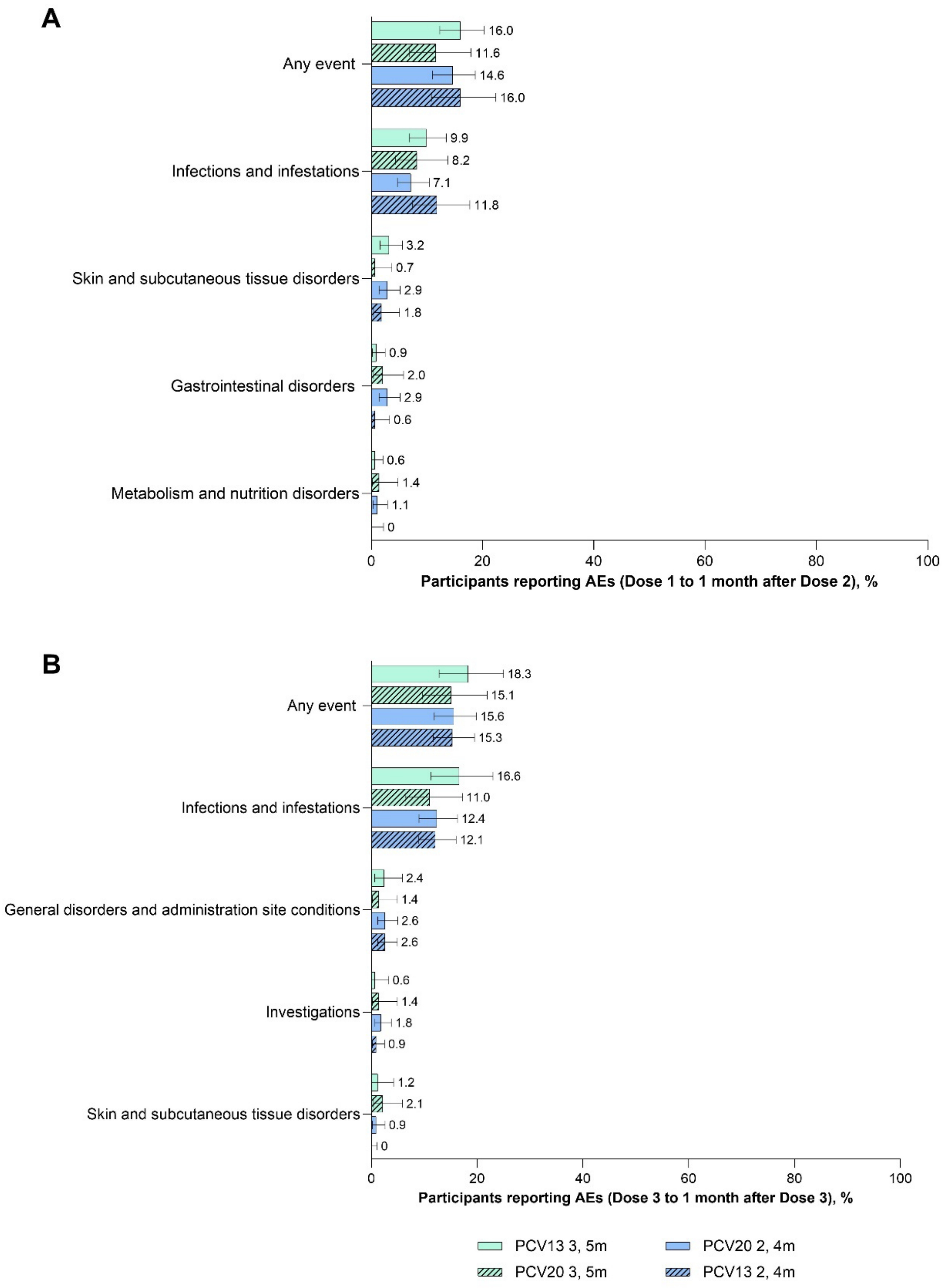

### Supplemental Content 6. Recommendations for maternal Tdap during pregnancy and PCV infant and toddler dose timings in Europe[1]

| Country | Maternal Tdap recommended during pregnancy | Recommended ages for PCV doses (months) |
|---|---|---|
| **2-, 4-month PCV schedules** | | |
| Belgium | Yes | 2, 4, 12 |
| Croatia | Yes | 2, 4, 12 |
| France | Yes | 2, 4, 11 |
| Germany | Yes | 2, 4, 11 |
| Greece | Yes | 2, 4, 12 |
| Hungary | Yes | 2, 4, 12 |
| Italy | Yes | 2, 4, 10 |
| Latvia | Yes | 2, 4, 12–15 |
| Liechtenstein | | 2, 4, 12 |
| Lithuania | Yes | 2, 4, 12–15 |
| Luxembourg | Yes | 2, 4, 11 |
| Malta | | 2, 4, 12 |
| Poland | Yes | 2, 4, 13–15 |
| Portugal | Yes | 2, 4, 12 |
| Romania | Yes | 2, 4, 11 |
| Spain | Yes | 2, 4, 11 |
| **3-, 5-month PCV schedules** | | |
| Austria | Yes | 3, 5, 12–14 |
| Cyprus | Yes | 3, 5, 12–15 |
| Denmark | Yes | 3, 5, 12 |
| Finland | | 3, 5, 12 |
| Iceland | Yes | 3, 5, 12 |
| Netherlands | Yes | 3, 5, 12 |
| Norway | | 3, 5, 12 |
| Slovakia | | 3, 5, 11 |
| Slovenia | Yes | 3, 5–6, 11–18 |
| Sweden | Yes | 3, 5, 12 |
| **Other PCV schedules** | | |
| Bulgaria | | 6 weeks, 3, 12 |
| Czechia | Yes | 2, 3–4, 5–6, 13–15 |
| Estonia | | No recommendations |
| Ireland | Yes | 2, 6, 13 |

PCV, pneumococcal conjugate vaccine; Tdap, tetanus, diphtheria, and pertussis vaccine.

[1]European Centre for Disease Prevention and Control Vaccine Scheduler. Vaccine schedules in all countries in the EU/EEA. Available at: https://vaccine-schedule.ecdc.europa.eu/ (accessed November 11, 2025).
